# Increased Hippocampal Blood Flow in People at Clinical High Risk for Psychosis and Effects of Cannabidiol

**DOI:** 10.1101/2023.02.22.23286306

**Authors:** Cathy Davies, Matthijs G Bossong, Daniel Martins, Robin Wilson, Elizabeth Appiah-Kusi, Grace Blest-Hopley, Fernando Zelaya, Paul Allen, Michael Brammer, Jesus Perez, Philip McGuire, Sagnik Bhattacharyya

## Abstract

**Background:** Hippocampal hyperperfusion has been observed in people at Clinical High Risk for Psychosis (CHR), is associated with adverse longitudinal outcomes and represents a potential treatment target for novel pharmacotherapies. Whether cannabidiol (CBD) has ameliorative effects on hippocampal blood flow (rCBF) in CHR patients remains unknown.

**Methods:** Using a double-blind, parallel-group design, 33 CHR patients were randomised to a single oral 600mg dose of CBD or placebo. Nineteen healthy controls were studied under identical conditions but did not receive any drug. Hippocampal rCBF was measured using Arterial Spin Labelling. We examined differences relating to CHR status (controls vs placebo), effects of CBD in CHR (placebo vs CBD) and linear between-group relationships, such that placebo>CBD>controls or controls>CBD>placebo, using a combination of hypothesis-driven and exploratory wholebrain analyses.

**Results:** Placebo-treated patients had significantly higher hippocampal rCBF bilaterally (all p_FWE_<.01) compared to controls. There were no suprathreshold effects in the CBD vs placebo contrast. However, we found a significant linear relationship in the right hippocampus (p_FWE_=.035) such that rCBF was highest in the placebo group, lowest in controls and intermediate in the CBD group. Exploratory wholebrain results replicated previous findings of hyperperfusion in the hippocampus, striatum and midbrain in CHR patients, and provided novel evidence of increased rCBF in inferior-temporal and lateral-occipital regions in patients under CBD compared to placebo.

**Conclusions:** These findings suggest that hippocampal blood flow is elevated in the CHR state and may be partially normalised by a single dose of CBD. CBD therefore merits further investigation as a potential novel treatment for this population.

## INTRODUCTION

Psychotic disorders such as schizophrenia are usually preceded by a prodromal stage, characterised by attenuated psychotic symptoms and social, emotional and cognitive dysfunction (Fusar-Poli *et al*., 2020b). Such individuals are said to be at Clinical High Risk for psychosis (CHR) and have a ∼22% three-year risk of transitioning to the full-blown disorder (Fusar-Poli *et al*., 2020b). The neurobiological mechanisms underlying psychosis risk and onset are incompletely understood (Millan *et al*., 2016) but compelling evidence implicates hippocampal dysfunction in its pathophysiology (Tamminga *et al*., 2010; Lieberman *et al*., 2018; Knight *et al*., 2022). Specifically, preclinical models suggest that hippocampal hyperactivity is key to the development of psychosis and arises due to NMDA receptor hypofunction on GABAergic interneurons, leading to disinhibition of hippocampal pyramidal cells (Lodge and Grace, 2007; Lisman *et al*., 2008) (Supplementary Fig S1). This is thought to drive hypermetabolism and elevated blood volumes/flow in the hippocampus (Schobel *et al*., 2013) as well as downstream midbrain-striatal hyperdopaminergia (Modinos *et al*., 2015; Grace and Gomes, 2019), with the consequent induction of psychotic-like phenotypes in animals and symptoms in humans (Fig S1).

These findings are consistent with *in vivo* evidence of increased cerebral blood volumes (CBV) in the hippocampus in patients with psychosis (Schobel *et al*., 2009; Talati *et al*., 2014, 2015; McHugo *et al*., 2019) and at baseline in CHR individuals who go on to transition (Schobel *et al*., 2009, 2013). However, numerous studies in patients with established illness have not found differences in hippocampal resting cerebral blood flow (rCBF, or ‘perfusion’) (for reviews see (Guimarães *et al*., 2016; Sukumar *et al*., 2020; Percie du Sert *et al*., 2023)). Although antipsychotics—which can impact rCBF (Goozée *et al*., 2014)—may be an important confounder here, few of the studies of antipsychotic-naïve or-free patients conducted to date (Medoff *et al*., 2001; Scheef *et al*., 2010; Selvaggi *et al*., 2022; Bojesen *et al*., 2023) report significant hippocampal rCBF differences.

By contrast, previous work in CHR patients demonstrates that rCBF in the hippocampus is increased relative to controls (Allen *et al*., 2016, 2018), with longitudinal reductions in hippocampal rCBF associated with remission from the CHR state (Allen *et al*., 2016). While only a minority of CHR individuals go on to develop frank psychosis (Fusar-Poli *et al*., 2020b), this clinical group offers a unique opportunity to investigate the mechanisms underlying psychosis risk without the effects of both antipsychotic exposure and illness chronicity. Hippocampal rCBF has also been associated with prefrontal GABA levels (Modinos *et al*., 2018b) and striatal dopamine function (Modinos *et al*., 2021) in CHR individuals, which are implicated in the final common pathways to psychosis (Howes and Kapur, 2009). Hippocampal hyperactivity (and associated hyperperfusion) may therefore represent potential treatment targets. Importantly, as the CHR state progresses to the first episode of psychosis, functional perturbations originating in the hippocampus appear to spread to regions such as the frontal cortex (Schobel *et al*., 2013; Lieberman *et al*., 2018), and excitotoxic as well as atrophic processes culminate in hippocampal volume loss (Ho *et al*., 2017b, 2017a; Vargas *et al*., 2017) and morphological alterations (Schobel *et al*., 2013) (Fig S1). If functional hippocampal changes precede structural (atrophic) changes, treatments targeted to hippocampal hyperactivity during the CHR stage may prove more effective as preventative strategies. However, there are currently no licensed pharmacological interventions for people at CHR (Davies *et al*., 2018a, 2018b), which remains a critical unmet need.

One of the most promising candidate treatments is cannabidiol (CBD), a phytocannabinoid constituent of the cannabis plant (Davies and Bhattacharyya, 2019). Compared to the main intoxicating cannabinoid in cannabis, delta-9-tetrahydrocannabinol (THC), which has psychotomimetic (D’Souza *et al*., 2004; Bhattacharyya *et al*., 2009; Morrison *et al*., 2009; Englund *et al*., 2013; Sherif *et al*., 2016) and potential anxiogenic effects, CBD is non-intoxicating and has anxiolytic (Bergamaschi *et al*., 2011; Crippa *et al*., 2011) and antipsychotic properties (Leweke *et al*., 2012; Crippa *et al*., 2018; McGuire *et al*., 2018). However, the mechanisms underlying these effects remain unclear. In healthy volunteers and patients with established psychosis, CBD modulates blood-oxygen-level-dependent (BOLD) haemodynamic responses to fMRI tasks in several regions, particularly medial temporal cortex and striatum, as well as functional connectivity between these regions (Fusar-Poli *et al*., 2009; Bhattacharyya *et al*., 2010, 2012, 2015; Gunasekera *et al*., 2020; O’Neill *et al*., 2021). In CHR patients, we previously demonstrated that a 600mg dose of CBD partially normalises mediotemporal and striatal function during verbal memory (Bhattacharyya *et al*., 2018) and fear processing (Davies *et al*., 2020), such that activation in the CBD group was intermediate between that of healthy controls and CHR patients under placebo. CBD has also been shown to modulate hippocampal perfusion. Two single-photon emission computed tomography (SPECT) studies found reductions in hippocampal blood flow following CBD in healthy individuals (Crippa *et al*., 2004) and patients with social anxiety disorder (Crippa *et al*., 2011). Conversely, a more recent study using Arterial Spin Labelling (ASL) found that CBD increased hippocampal rCBF in healthy participants, a specific effect not found in five other regions-of-interest (ROIs) (Bloomfield *et al*., 2019). The presence of hippocampal effects across all three prior studies is encouraging and suggests that CBD may engage one of the most strongly implicated neurobiological treatment targets for people at CHR. However, whether CBD has ameliorative effects on hippocampal blood flow in CHR patients remains to be investigated.

To address this gap, in the present study we used ASL to examine hippocampal blood flow in three parallel groups: CHR patients randomised to a single 600mg dose of CBD or placebo and healthy controls. On the basis of data from previous studies in CHR populations (Allen *et al*., 2016, 2018), we selected bilateral ROIs within the hippocampus and assessed extra-hippocampal effects with exploratory wholebrain analyses. We first examined whether CHR patients under placebo conditions show elevated hippocampal blood flow compared to healthy controls. We then tested our primary hypothesis that CHR patients receiving CBD would show at least partial ‘normalisation’ of hippocampal blood flow in the same regions identified as different in the placebo vs control analyses. That is, perfusion in the CBD group would be intermediate between that observed in healthy controls and the CHR placebo group.

## PATIENTS & METHODS

### Participants

The study was registered (ISRCTN46322781) and received Research Ethics (Camberwell St Giles) approval. All participants provided written informed consent. Thirty-three antipsychotic-naive CHR individuals, aged 18–35, were recruited from specialist early detection services in the United Kingdom. CHR status was determined using the Comprehensive Assessment of At-Risk Mental States (CAARMS) criteria (Yung *et al*., 2005). Briefly, subjects met one or more of the following subgroup criteria: (a) attenuated psychotic symptoms, (b) brief limited intermittent psychotic symptoms (psychotic episode lasting <1 week, remitting without treatment), or (c) either schizotypal personality disorder or first-degree relative with psychosis, all coupled with functional decline (Yung *et al*., 2005). Nineteen age (within 3 years), sex and ethnicity-matched healthy controls were recruited locally by advertisement. Exclusion criteria included history of psychotic or manic episode, current DSM-IV diagnosis of substance dependence (except cannabis), IQ<70, neurological disorder or severe intercurrent illness, and any contraindication to magnetic resonance imaging (MRI) or treatment with CBD. Participants were required to abstain from cannabis for 96h, other recreational substances for 2 weeks, alcohol for 24h and caffeine and nicotine for 6h before attending. A urine sample prior to scanning was used to screen for illicit drug use and pregnancy.

### Design, Materials, Procedure

Using a randomised, double-blind, placebo-controlled, three-arm parallel-group design, CHR participants were randomised to a single oral 600mg dose of CBD (THC-Pharm) or a matched placebo capsule. This dose was selected based on previous findings that doses of 600-800 mg/day are effective in established psychosis (Leweke *et al*., 2012) and anxiety (Bergamaschi *et al*., 2011). Psychopathology was measured at baseline (before drug administration) using the CAARMS (positive and negative symptoms) and State-Trait Anxiety Inventory (State Subscale). Following a standard light breakfast, participants were administered the capsule (at ∼11AM) and 180 min later, underwent a battery of MRI sequences. This interval between drug administration and MRI acquisition was selected based on previous findings describing peak plasma concentrations at 180min following oral administration (Martin-Santos *et al*., 2012; Millar *et al*., 2018). Control participants were investigated under identical conditions but did not receive any drug. Plasma CBD levels were sampled at baseline and at 120 and 300 min after drug administration.

### MRI Acquisition and Image Processing

All scans were acquired on a General Electric Signa HDx 3T MR system with an 8-channel head coil. For image registration, high resolution T2-weighted Fast Spin Echo (FSE) and T1-weighted Spoiled Gradient Recalled images were acquired. Resting CBF was measured using pseudo-Continuous ASL acquired with a 3D-FSE spiral multi-shot readout. Acquisition parameters and preprocessing procedures (conducted using FMRIB Software Library; FSL/6.0.2) were in line with previous studies (Allen *et al*., 2016, 2018; Modinos *et al*., 2018b, 2021) and are detailed in the Supplementary Methods.

### Statistical Analysis

#### Global Blood Flow

To exclude potential group differences in global CBF, we extracted mean CBF values from the MNI152 grey matter mask (thresholded at >.50 probability) for each subject using *fslmeants* in FSL. We then conducted analyses of covariance (ANCOVA) in SPSS 27 using mean-centred age, sex, education and smoking as covariates. All subsequent analyses were conducted with global CBF as a nuisance covariate.

#### Hippocampal Blood Flow

Analyses of rCBF data were conducted using Statistical Parametric Mapping 12 (SPM12) in Matlab/R2018b using an ROI approach. Hippocampal ROIs were specified *a priori* using coordinates from a previous (Allen *et al*., 2016) (and replicated (Allen *et al*., 2018)) study comparing CBF in CHR patients vs healthy controls, which have since been used in studies across the extended psychosis phenotype (Modinos *et al*., 2018b, 2018a); MNI coordinates in right: x=20, y=−28, z=−8 and left hippocampus: x=−22, y=−28, z=−8. 6mm spheres around these coordinates (123 voxels per ROI) were combined into a single volume and used as an explicit mask (246 voxels in total). In line with our first objective, we used an independent samples t-test to compare the placebo-treated CHR group with healthy controls to identify differences in rCBF (within our pre-defined ROIs) related to CHR status. In a second independent samples t-test, we directly compared CHR patients under placebo with those under CBD to test whether CBD had effects on rCBF in the same ROIs. Finally, to test our primary hypothesis that rCBF in the CBD group would be intermediate between that of the healthy control and placebo groups (which may suggest partial normalisation of rCBF by CBD), we examined whether a linear relationship in rCBF (controls>CBD>placebo; or placebo>CBD>controls) existed within the same ROIs using ANCOVA (flexible factorial). Linear trend (i.e. linear relationship) analyses are distinct from standard ANOVA in that they test for specific relationships (such as linear or quadratic) across groups, which are not tested by the standard F-test (Howell, 2010). In line with previous CHR studies of CBF (Allen *et al*., 2016, 2018), mean-centred age, sex, smoking status and years of education (the latter included due to significant group differences in our sample) were included in all analyses as nuisance covariates, as was mean global CBF (via global normalisation in SPM). Results were considered significant after p<.05 with family-wise error (FWE) correction for multiple comparisons at the voxel level, as in previous CHR studies (Allen *et al*., 2018; Modinos *et al*., 2018a).

#### Exploratory Wholebrain Analyses

For completeness, we examined the same contrasts as above at wholebrain level. We conducted a wholebrain search using the explicit grey matter mask (again thresholded at >.50) and cluster-level inference (cluster-forming threshold: p<.005; cluster reported as significant at p<.05 using FWE cluster correction in SPM). Supplemental analyses to evaluate the robustness of significant wholebrain findings were conducted using FSL’s randomise, as detailed in the Supplementary Material.

#### Demographics

Analyses of baseline and demographic variables were conducted in SPSS using independent t-tests for continuous data and chi-square tests for categorical data. Significance was set at p<.05.

## RESULTS

There were no between-group differences in the majority of demographic and baseline clinical characteristics, except for fewer years of education in the placebo group relative to controls (Table 1), as reported in our previous publications (Bhattacharyya *et al*., 2018; Wilson *et al*., 2019; Davies *et al*., 2020). In the CBD group, mean (SD) plasma CBD levels were 126.4nM (221.8) and 823.0nM (881.5) at 120 and 300 min after drug intake, respectively (Supplementary Fig S3). Three CHR individuals exited the scanner prior to the ASL sequence, and two CHR subjects’ data were corrupted at source, leaving n=14 in the placebo group, n=14 in the CBD group and n=19 controls (see Supplementary CONSORT details).

**TABLE 1.** Total Sample Sociodemographic and Clinical Characteristics.

| Characteristic |  | CBD<br>(n = 16) | Placebo<br>(n = 17) | Control<br>(n = 19) <sup>1</sup> | Pairwise Comparison |  |
| --- | --- | --- | --- | --- | --- | --- |
|  |  |  |  |  | Control vs<br>Placebo | Placebo<br>vs CBD |
| Age, years; mean (SD) |  | 22.7 (5.08) | 24.1 (4.48) | 23.9 (4.15) | p=.91 <sup>2</sup> | p=.42 <sup>2</sup> |
| Sex, N (%) male |  | 10 (62.5) | 7 (41.2) | 11 (57.9) | p=.32 <sup>3</sup> | p=.22 <sup>3</sup> |
| Ethnicity,<br>N (%) | White | 10 (62.5) | 7 (41.2) | 11 (57.9) | p=.59 <sup>3</sup> | p=.43 <sup>3</sup> |
|  | Black | 2 (12.5) | 5 (29.4) | 5 (26.3) |  |  |
|  | Asian | 0 (0) | 1 (5.9) | 0 (0) |  |  |
|  | Mixed | 4 (25) | 4 (23.5) | 3 (15.8) |  |  |
| Education, years; mean (SD) |  | 14.4 (2.71) | 12.6 (2.76) | 16.9 (1.58) | p<.001 <sup>2</sup> | p=.06 <sup>2</sup> |
| Handedness, N (%) right |  | 14 (87.5) | 17 (100) | 18 (94.7) | p=.37 <sup>3</sup> | p=.16 <sup>3</sup> |
| CAARMS Positive, mean (SD) |  | 40.19 (20.80) | 42.94 (29.47) | NA | NA | p=.76 <sup>2</sup> |
| CAARMS Negative, mean (SD) |  | 23.25 (16.49) | 28.41 (20.49) | NA | NA | p=.43 <sup>2</sup> |
| STAI-S, mean (SD) |  | 40.31 (9.07) | 38.94 (10.18) | NA | NA | p=.69 <sup>2</sup> |
| Urine drug<br>screen<br>results,<br>N (%) | Clean | 10 (63) | 8 (47) | NA <sup>4</sup> | Not<br>compared <sup>1</sup> | p=.45 <sup>3</sup> |
|  | THC | 2 (13) | 5 (29) | NA <sup>4</sup> |  |  |
|  | Morphine | 1 (6) | 0 (0) | NA <sup>4</sup> |  |  |
|  | Benzodiazepines | 0 (0) | 1 (6) | NA <sup>4</sup> |  |  |
|  | PCP | 0 (0) | 1 (6) | NA <sup>4</sup> |  |  |
|  | Missing | 3 (19) | 2 (12) | NA <sup>4</sup> |  |  |
| Current nicotine use, N (%) yes |  | 9 (56) | 5 (29) | 2 (11) | p=.15 <sup>3</sup> | p=.12 <sup>3</sup> |
| Current alcohol use, N (%) yes |  | 11 (69) | 10 (59) | NA | Not<br>compared <sup>1</sup> | p=.59 <sup>3</sup> |
| Lifetime cannabis use, N (%)<br>yes |  | 15 (94) | 17 (100) | NA <sup>5</sup> | Not<br>compared <sup>1</sup> | p=.48 <sup>3</sup> |
| Current cannabis use, N (%) yes |  | 7 (44) | 7 (41) | NA <sup>5</sup> | Not<br>compared <sup>1</sup> | p=.88 <sup>3</sup> |
| Cannabis<br>use<br>frequency,<br>N (%)<sup>6</sup> | More than once a<br>week | 11 (69) | 12 (71) | NA | Not<br>compared <sup>1</sup> | p=.38 <sup>3</sup> |
|  | Once/twice<br>monthly | 1 (6) | 3 (18) | NA |  |  |
|  | Few times a year | 2 (13) | 0 (0) | NA |  |  |
|  | Only once/twice<br>lifetime | 1 (6) | 2 (12) | NA |  |  |

**Abbreviations:** CAARMS, Comprehensive Assessment of At-Risk Mental States; CBD, cannabidiol; CHR, Clinical High Risk for Psychosis; N, number of subjects; NA, not applicable; PCP, phencyclidine; STAI-S, State-Trait Anxiety Inventory-State Subscale; THC, $\Delta$ 9-tetrahydrocannabinol. <sup>1</sup>Controls were selected to have minimal drug use and hence were not compared with CHR participants on these parameters; <sup>2</sup>Independent t-test; <sup>3</sup>Pearson chi-squared test; <sup>4</sup>Controls tested negative on urine drug screen for all substances tested; <sup>5</sup>Cannabis use less than 10 times lifetime (no current users); <sup>6</sup>Note that this data are reported for all subjects who had ever used cannabis, regardless of whether they were current users.

### Global CBF

There were no significant group differences in mean global grey matter CBF values (ml/100g/min): healthy control (marginal mean ± SE)= 48.96 ± 2.92; CBD= 45.39 ± 2.88; placebo= 47.79 ± 3.27; F(2,40)=0.38, p=.69.

### Hippocampal Blood Flow – Pairwise effects of CHR status and CBD

Pairwise t-tests examining differences related to CHR status revealed that compared to healthy controls, placebo-treated patients had significantly greater rCBF in the hippocampus bilaterally (right hippocampus: MNI coordinates X=22 Y=-24 Z=-4, T(26)=5.29, Z_E_=4.32, k=90, p<.001 FWE; left hippocampus: MNI coordinates X=-24 Y=-24 Z=-10, T(26)=4.01, Z_E_=3.50, k=96, p=.009 FWE; Fig 1A). There were no suprathreshold voxels in the CBD vs placebo pairwise comparison.

**FIGURE 1.**
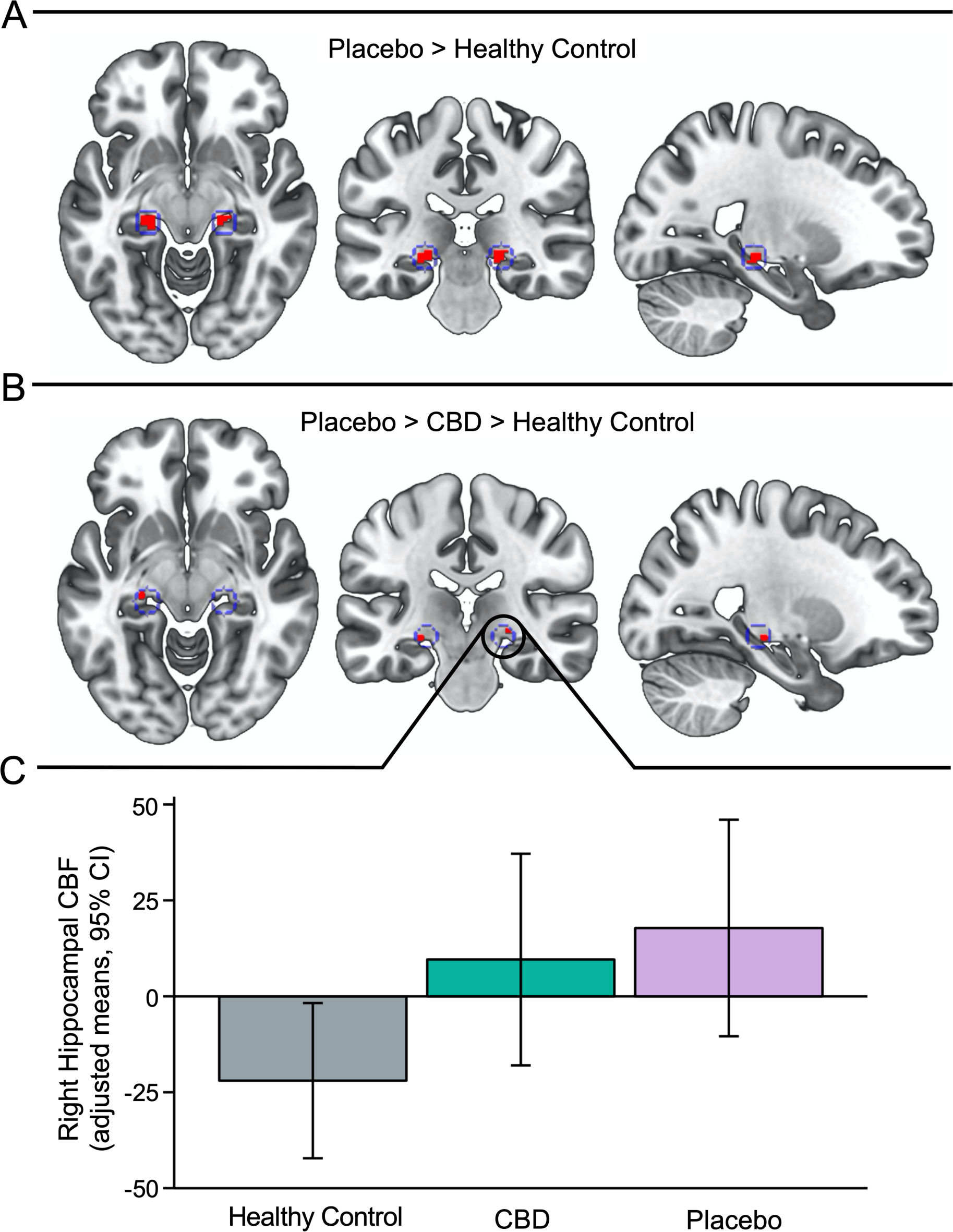
Hippocampal blood flow across CHR patients receiving placebo, CHR patients receiving CBD and healthy controls. **(A)** Increased blood flow (shown in red) in CHR patients in the placebo group relative to healthy controls in right (p<.001 FWE) and left hippocampus (p=.009 FWE), overlaid on a standard brain template. Hippocampal ROIs are depicted by the inner bounds of the blue circles. **(B)** Voxels where blood flow followed a linear pattern across groups (placebo > CBD > controls) in right (p=.035 FWE) and left hippocampus (p=.053 FWE), overlaid on a standard brain template. Hippocampal ROIs are depicted by the inner bounds of the blue circles. The right side of the brain is shown on the right of the axial and coronal images. The left side of the brain is shown in the sagittal images. **(C)** Plot showing group means ± 95% CIs of the covariate-adjusted mean rCBF values for the right hippocampal cluster (indicated in panel B) as extracted from the F-test design matrix for each subject using the MarsBaR toolbox in SPM (arbitrary units). The adjusted means ± SD (arbitrary units) were: placebo (M= 17.81, SD= 48.9), CBD (M= 9.59, SD= 47.8), and healthy controls (M= −21.98, SD= 42.0).

### Hippocampal Blood Flow – Between-Group Linear Analyses

Our primary *a priori* analysis revealed a significant linear relationship in the right hippocampus, such that blood flow was highest in the placebo group, lowest in healthy controls, and intermediate in the CBD group (MNI coordinates X=24 Y=-24 Z=-6, F(2,39)=8.01, Z_E_=3.03, k=4, p=.035 FWE; Fig 1B-C). The relationship in the left hippocampus was not significant but followed the same pattern (placebo>CBD>controls) at trend-level (X=-26 Y=-24 Z=-10, F(2,39)=7.30, Z_E_=2.87, k=8, p=.053 FWE; Fig 1B).

### Exploratory Wholebrain Analyses

In pairwise analyses for main effect of CHR status, compared to healthy controls, placebo-treated patients had significantly higher CBF in two clusters spanning parts of the bilateral hippocampus, parahippocampal gyrus, midbrain/brainstem, thalamus, cerebellum and left striatum (mostly putamen) and amygdala (2 clusters, both p<.01 FWE_cluster_; Fig 2). In pairwise analyses for the main effect of CBD in CHR, we found significantly higher CBF in left inferior and middle temporal gyri (temporo-occipital parts) and lateral occipital regions in the CBD compared to the placebo group (p=.014 FWE_cluster_; Fig 2). There were no suprathreshold clusters in the wholebrain three-group linear analyses. Supplementary analyses conducted to test the robustness of these findings (using FSL’s *randomise* rather than SPM) showed almost identical significant results for the placebo > control contrast. However, while the cluster found in the CBD > placebo contrast (above) was present at a relaxed statistical threshold, it was not significantly different between the groups (p=.15 FWE_cluster_; see Supplementary Material).

**FIGURE 2.**
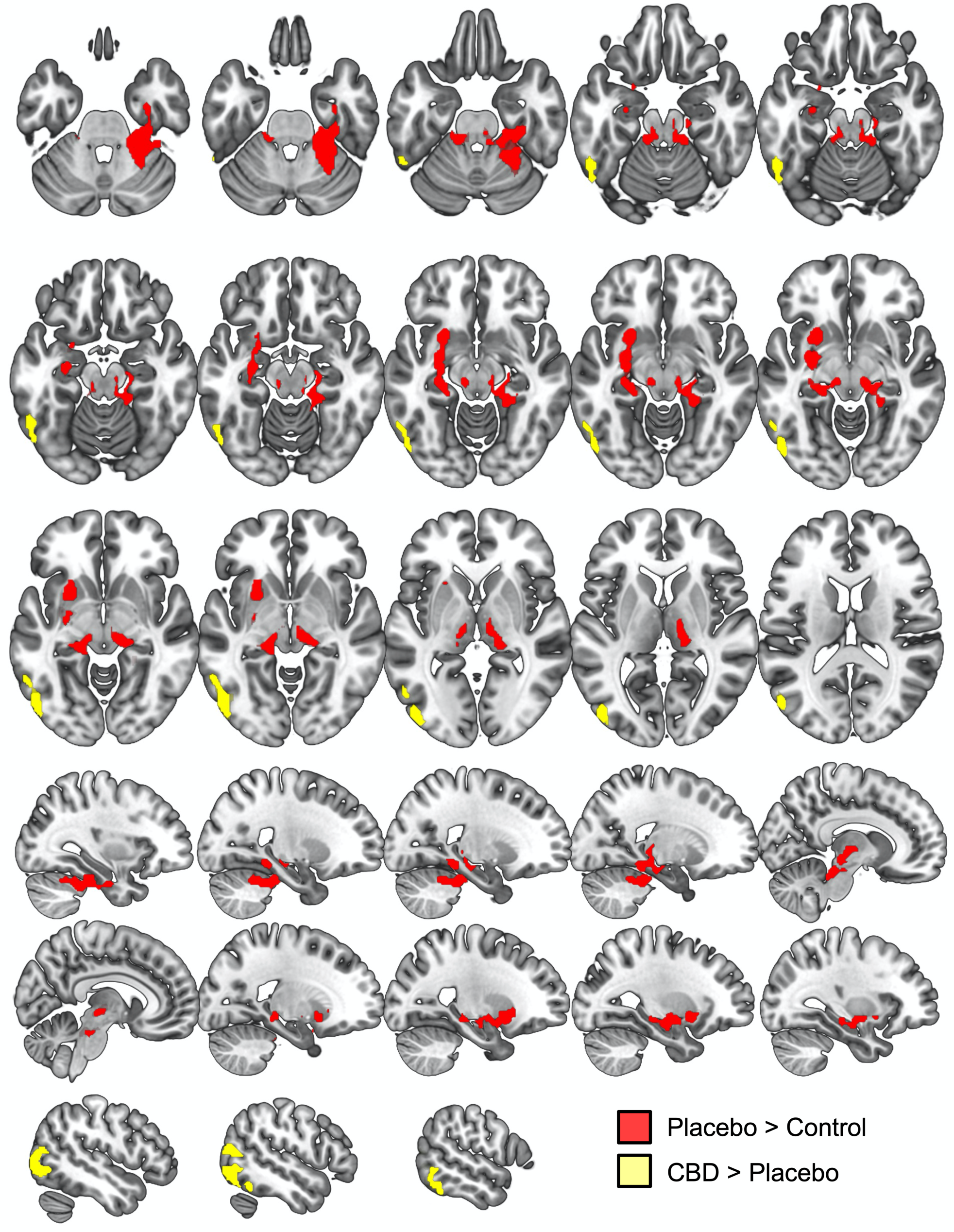
Differences in CBF related to CHR status (placebo vs healthy controls) and effects of CBD in CHR patients (CBD vs placebo) from wholebrain analyses. Axial and sagittal sections showing significant clusters of group differences in CBF from wholebrain analyses. Two large significant clusters were identified for the placebo > control contrast (regions shown in red). Cluster 1 spanned portions of the left hippocampus, parahippocampal gyrus, amygdala, midbrain/brainstem, thalamus, putamen, pallidum and cerebellum (cluster 1 peak MNI coordinates X= −26 Y= 10 Z= −8, T(26)= 5.17, ZE= 4.25, k=735, p=.008 FWEcluster). Cluster 2 spanned portions of the right thalamus, parahippocampal gyrus, lingual gyrus, temporal fusiform cortex, cerebellum, midbrain/brainstem and hippocampus (cluster 2 peak MNI coordinates X= 20 Y= - 22 Z= −16, T(26)= 4.85, ZE= 4.05, k=1271, p<.001 FWEcluster). One significant cluster was identified in the CBD > placebo analysis (regions shown in yellow), which spanned portions of the left lateral occipital cortex (inferior division) and middle and inferior temporal gyri (temporo-occipital parts) (peak MNI coordinates X= −44 Y= −80 Z= 6, T(21)= 4.51, ZE= 3.75, k=638, p=.014 FWEcluster). There were no significant results for the control > placebo contrast, the placebo > CBD contrast, nor either of the three-way linear contrasts. The right side of the brain is shown on the right of the axial images. The sagittal sections are ordered from right to left hemispheres (top row to bottom row).

## DISCUSSION

This is the first study to investigate the effects of CBD on cerebral blood flow in patients at CHR for psychosis. We first established that hippocampal rCBF is elevated in CHR patients under placebo compared to healthy controls. To test our primary hypothesis that CBD would at least partially normalise any such alterations in hippocampal rCBF, we then examined whether a linear relationship existed between the groups. In line with our predictions, our key finding was that hippocampal blood flow was indeed highest in the placebo-treated CHR group, lowest in healthy controls and significantly intermediate in the CBD-treated CHR group. Together, these findings provide the first—albeit preliminary—*in vivo* evidence that CBD may engage and partially normalise one of the key neurobiological treatment targets for patients at risk of psychosis. Given the current lack of effective pharmacotherapies for CHR patients (Davies *et al*., 2018a, 2018b), CBD represents a promising compound for future preventive studies.

### Hippocampal Blood Flow is Elevated in CHR Patients

Our first key finding was that hippocampal blood flow is increased in CHR (placebo) patients vs controls. This is consistent with studies to have addressed this issue previously (Allen *et al*., 2016, 2018) as well as contemporary models suggesting that hippocampal hyperactivity is present prior to psychosis onset and plays an upstream role in its progression (Schobel *et al*., 2013; Lieberman *et al*., 2018). In using ROIs based on hippocampal clusters previously found to be altered in CHR patients (Allen *et al*., 2016, 2018), our results provide a further external replication of elevated rCBF in these regions in an independent CHR sample. Notably, in line with prior work (Schobel *et al*., 2013; Allen *et al*., 2016) we found hyperperfusion in the hippocampus bilaterally, whereas some previous studies using the same hippocampal ROIs find effects only on the right side, both in CHR (Allen *et al*., 2018) and high schizotypy (Modinos *et al*., 2018a) samples.

However, despite substantial evidence of elevated CBV in the hippocampus (and/or specific subfields, particularly CA1) in psychosis (Schobel *et al*., 2009, 2013; Talati *et al*., 2014, 2015; McHugo *et al*., 2019), elevated hippocampal rCBF is not always found *in vivo* in patients with first-episode and/or established illness (Horn *et al*., 2009; Scheef *et al*., 2010; Pinkham *et al*., 2011; Walther *et al*., 2011; Ota *et al*., 2014; Talati *et al*., 2015; Bojesen *et al*., 2023) (for reviews and meta-analyses see (Guimarães *et al*., 2016; Sukumar *et al*., 2020; Percie du Sert *et al*., 2023)). Why elevated hippocampal rCBF would be more readily observed—so far, at least— in studies of patients at CHR rather than with established psychosis remains unclear. One potential contributing factor is that antipsychotics impact rCBF (Goozée *et al*., 2014)—but not CBV (Schobel *et al*., 2009)—and although this is typically reported within basal ganglia (Hawkins *et al*., 2017; Bojesen *et al*., 2023), effects are also seen in hippocampus (Medoff *et al*., 2001; Lahti *et al*., 2003). However, several studies have investigated rCBF in antipsychotic-free or-naïve patients (Medoff *et al*., 2001; Scheef *et al*., 2010; Selvaggi *et al*., 2022; Bojesen *et al*., 2023) and few (Medoff *et al*., 2001) report significant hippocampal differences. Another possibility is that the cascading pathophysiology is illness stage-specific and evolves over the course of psychosis progression (Millan *et al*., 2016). This would be consistent with evidence that functional changes (observed in the CHR state) precede structural changes (seen after psychosis onset and in those who do not remit), with initial CA1 hypermetabolism spreading to other subfields and extra-hippocampal regions (such as frontal cortex) in those who transition, before hippocampal volume loss and morphological changes become apparent (Schobel *et al*., 2009, 2013; Ho *et al*., 2017b, 2017a). Overall, several studies (including the present one) now show that hippocampal rCBF is elevated in people at CHR (Allen *et al*., 2016, 2018), a patient group where important factors such as antipsychotic exposure and illness chronicity (Kraguljac and Lahti, 2021) are minimal. However, it should be noted that the majority of patients meeting CHR criteria (78% at 3 years, 63% at 10-11 years (Fusar-Poli *et al*., 2020b, 2020a)) do not go on to develop psychosis. As such, further research measuring rCBF and CBV longitudinally over the course of psychosis progression is required to unravel the precise nature, temporal sequence and relevance of these findings to the mechanisms of psychosis risk vs frank illness.

### CBD May Attenuate Hippocampal Blood Flow

Our second key finding—that hippocampal blood flow in CBD-treated CHR patients is intermediate between controls and placebo-treated patients—is translationally relevant, given previous evidence (above) linking hippocampal hypermetabolism in the CHR state with the onset of psychosis (Schobel *et al*., 2013) and longitudinal reductions in hippocampal CBF associating with CHR remission (Allen *et al*., 2016). The reduction of hippocampal perfusion (as suggested here) would thus seem a plausible therapeutic target for novel antipsychotic treatments. Elevated hippocampal rCBF and blood volumes are also positively associated with symptom severity across psychosis and CHR patients (Schobel *et al*., 2009), as well as delusional thinking in non-help-seeking healthy individuals (Wolthusen *et al*., 2018). A partial normalisation of hippocampal rCBF by CBD could, therefore, underlie or contribute to the therapeutic effects of CBD that have been reported in patients with established psychosis (Leweke *et al*., 2012; McGuire *et al*., 2018) (see Supplementary Material for discussion of potential mechanisms). However, our cross-sectional study was designed and powered to examine acute neurophysiological rather than symptom effects and as such, future longitudinal studies administering CBD to a larger sample are needed to evaluate clinical efficacy.

Our findings are consistent with previous literature on the effects of CBD. Using the same patient and control sample, we previously demonstrated that CBD has effects on task-based BOLD haemodynamic readouts (Bhattacharyya *et al*., 2018; Wilson *et al*., 2019; Davies *et al*., 2020), finding the same commensurate pattern of placebo > CBD > controls (or vice versa) in mediotemporal regions during fear processing and verbal memory fMRI (Bhattacharyya *et al*., 2018; Davies *et al*., 2020). CBF is intrinsically linked to BOLD responses via neurovascular coupling (Kim *et al*., 2020), but its acquisition does not require the cognitive or other manipulation needed for task-based fMRI contrasts (Alsop *et al*., 2015). As such, our results extend previous work by suggesting that CBD may also attenuate basal resting-state hippocampal activity in CHR patients (see Supplementary Discussion). Albeit in the context of our linear between-group results, the direction of CBD effects we observed is also in line with two previous SPECT studies, which found significant reductions in hippocampal rCBF following CBD in healthy people and those with anxiety disorders (Crippa *et al*., 2004, 2011). Conversely, the direction of our effects contrasts with the only other study to have examined CBD using ASL, which found a significant increase in hippocampal rCBF in healthy individuals (Bloomfield *et al*., 2019). In our view, the most likely explanation for this difference is that the effects of CBD differ as a function of the sample population and more specifically, the presence vs absence of baseline pathophysiology. Several lines of disparate evidence support this notion. For example, CBD has been shown to have different effects on reward processing-related activity (Wilson *et al*., 2019; Lawn *et al*., 2020; Gunasekera *et al*., 2022), GABA levels (Pretzsch *et al*., 2019a) and resting-state brain activity (Pretzsch *et al*., 2019b) across various healthy and clinical populations. Collectively, these findings suggest that the direction of CBD’s effects may depend on the specific clinical vs healthy population under study, with CBD potentially having effects in the direction towards normalisation in those with baseline dysfunction. A broader implication is that if effects in clinical populations cannot accurately be extrapolated from findings in healthy cohorts, future pharmacological studies to establish group-specific target engagement remain of particular value.

Although we found significant effects in our *a priori* regions and in the direction that appears to be of therapeutic and translational relevance, it is worth noting that the resulting cluster of voxels (with statistics based on voxel peak) in the linear relationship analyses was small (k=4), and we did not find any suprathreshold effects in the direct CBD vs placebo pairwise analysis. A significant cluster with greater spatial extent, and significant CBD vs placebo pairwise differences, would have provided more convincing evidence of CBD effects, and thus we await future replication and confirmation/refutation of these findings in future studies with larger sample sizes. However, regarding the latter point, the rationale for performing the three-way linear analyses—as implemented in our previous publications (Bhattacharyya *et al*., 2018; Wilson *et al*., 2019; Davies *et al*., 2020)—was that we do not necessarily expect that a single dose of CBD will fully reverse aberrant rCBF in CHR patients. Rather, we hypothesised that a signal of change towards normalisation in the CBD group (i.e. a significant linear relationship, suggestive of ‘partial normalisation’ effects) would provide initial evidence of disease-target engagement, in a direction indicative of therapeutic effects. Based on our initial findings here, future studies administering CBD repeatedly over longer durations are warranted.

### Exploratory Wholebrain Results

Outside of the hippocampus, in exploratory wholebrain analyses we found significant hyperperfusion in CHR (placebo) patients compared to controls in each of the major nodes implicated in psychosis pathophysiology by preclinical models and extant literature (Lodge and Grace, 2011; Grace and Gomes, 2019). This included the striatum, midbrain, thalamus and cerebellum, as well as bilateral hippocampi. Very few CHR perfusion studies have utilised a wholebrain approach, but one study reported corresponding hyperperfusion in most of these regions (Allen *et al*., 2016), although another study did not (Kindler *et al*., 2018). Effects in these regions also correspond with results of previous ROI-based studies, which include increased putamen rCBF in CHR patients (Kindler *et al*., 2018) which was correlated with positive symptom severity. In the same sample as the present study, we recently combined hippocampal glutamate data with wholebrain CBF maps and found an atypical relationship between glutamate and striatal-insula perfusion in CHR-placebo patients relative to controls (Davies *et al*., 2023), suggesting that striatal perfusion abnormalities may be linked to aberrant hippocampal neurochemistry. Greater perfusion in the putamen and thalamus has also been identified as a potential marker of genetic susceptibility for schizophrenia spectrum disorders in a neuroimaging twin study (Legind *et al*., 2019). Conversely, although both increased (Allen *et al*., 2016) and decreased (Kindler *et al*., 2018) prefrontal perfusion has been observed in CHR and established psychosis groups (Schobel *et al*., 2009), we did not find differences here in the present exploratory analyses.

In terms of wholebrain CBD effects, we found significantly increased CBF in inferior-temporal and lateral-occipital regions in CBD-treated relative to placebo-treated CHR patients. A previous SPECT study in healthy individuals reported attenuated perfusion in medial-occipital and inferior-temporal regions following CBD, albeit at a relaxed significance threshold (Crippa *et al*., 2004). The exploratory nature of our findings here combined with the paucity of prior literature means that their relevance is uncertain. Moreover, in view of the potential divergent effects of CBD on CBF across samples, these wholebrain effects may well differ in healthy or other clinical populations.

### Limitations

In addition to the limited spatial extent of the cluster identified in our primary (linear relationship) results, several further limitations warrant consideration. The first is that our study was parallel-group rather than within-subject. Although we used a randomised, double-blind design and there were no baseline differences between the two CHR groups (including in measures of substance use), the possibility that any between-group differences we observed were attributable to between-subject variability, as opposed to an effect of CBD, cannot be completely excluded. Future work administering both placebo and CBD in the same CHR (and healthy) individuals would address this issue and permit full factorial analyses. Such a design would also rule out potential expectation effects in the CHR placebo group in the contrast with controls. In terms of our patient group, we recruited a representative sample of CHR individuals as typically found in specialist CHR services (Fusar-Poli *et al*., 2020c). However, the CHR population is clinically heterogeneous (Fusar-Poli *et al*., 2020b) and thus the effects of CBD may be greater (or different) in specific subgroups of patients. Future studies with larger sample sizes are needed to stratify results by transition status or, for example, the three component subgroups of the CAARMS (Fusar-Poli *et al*., 2020b). Finally, this study reports on the acute neurophysiological effects of CBD and it is possible that the effects may differ after a sustained period of treatment. Future work by our group aims to address this issue while examining the effects of CBD on psychotic symptoms.

### Conclusion

Our findings indicate that a single dose of CBD may partially normalise aberrant hippocampal perfusion in CHR patients, a potential pathophysiological marker implicated in psychosis risk. Moreover, this effect occurred in the same region we show to be altered in CHR patients under placebo relative to healthy controls. CBD therefore merits further investigation as a candidate novel treatment for this group.

## ETHICAL STANDARDS

The authors assert that all procedures contributing to this work comply with the ethical standards of the relevant national and institutional committees on human experimentation and with the Helsinki Declaration of 1975, as revised in 2008.

## Data Availability

Data produced in the present study are not openly available.

## ACKNOWLEDGEMENTS

The authors wish to thank the study volunteers for their participation, Brandon Gunasekera (IoPPN, King’s College London) for his technical advice, and the radiographers at the Centre for Neuroimaging Sciences, King’s College London, who carried out the MRI scans.

## AUTHOR CONTRIBUTIONS

Substantial contributions to conception and design (SB, PMG, PA, MGB, FZ), acquisition of data (RW, EAK, GBH), analysis and/or interpretation of data (CD, SB, DM, MGB, FZ), drafting of the article (CD, SB) or revising it critically for important intellectual content (all authors), study supervision (SB). Final approval of the version to be published: all authors. CD and SB agree to be accountable for all aspects of the work in ensuring that questions related to the accuracy or integrity of any part of the work are appropriately investigated and resolved.

## PRE-PRINT

A pre-print version of this manuscript was deposited on the medRxiv pre-print server: https://doi.org/10.1101/2023.02.22.23286306.

## OPEN ACCESS

For the purposes of open access, the author has applied a Creative Commons Attribution (CC BY) licence to any Accepted Author Manuscript version arising from this submission.

