## Supplementary Material for "Increased Hippocampal Blood Flow in People at Clinical High Risk for Psychosis and Effects of Cannabidiol"

**Supplementary Introduction**
- Fig S1. Schematic of neural circuit mechanisms of hippocampal dysfunction in the pathophysiology underlying psychosis onset

**Supplementary Methods**
- MRI Acquisition
- MRI Processing
- SPM Thresholds

**Supplementary Results**
- CONSORT Details
- Fig S2. CONSORT Flow Diagram
- Fig S3. CBD Plasma Levels
- Supplementary Wholebrain Analyses: SPM vs FSL Randomise
- Exploratory Correlations between CAARMS and rCBF

**Supplementary Discussion**

**Supplementary References**

**FIGURE S1. Schematic of proposed neural circuit mechanisms of hippocampal dysfunction in the pathophysiology underlying psychosis onset.** In (1), low glutamate signal/input from hypofunctioning NMDARs (akin to faulty homeostatic sensors) prompts GABAergic interneurons to homeostatically increase excitation by reducing inhibition (disinhibition) of glutamatergic pyramidal cells. However, by disinhibiting pyramidal cells (and thus increasing glutamate signalling) in this dysfunctional neural environment, the potential homeostatic adaptation becomes allostatic, with enhanced excitatory drive inducing (2) hypermetabolism and hyperperfusion (elevated blood flow to meet increased metabolic demand), and (3) an overdrive in the responsivity of midbrain dopamine neurons, which project to the associative striatum. Note that the connection between hippocampal pyramidal cells and midbrain dopamine neurons is presented as monosynaptic but is in fact polysynaptic via the ventral striatum and ventral pallidum. Completing the (simplified) circuit, local glutamatergic tone is increased in (4) but is not detected as such by hypofunctioning NMDARs on GABAergic interneurons. Figure reproduced and adapted with permission (CCBY 4.0) from (Davies *et al.*, 2019). For original diagrams and discussion of evidence for this proposed circuit, see (Lisman *et al.*, 2008; Krystal and Anticevic, 2015; Modinos *et al.*, 2015; Krystal *et al.*, 2017; Lieberman *et al.*, 2018). *Abbreviations:* Glu, glutamate; NMDAR, N-methyl-D-aspartate receptor; CA1, Cornu Ammonis 1.

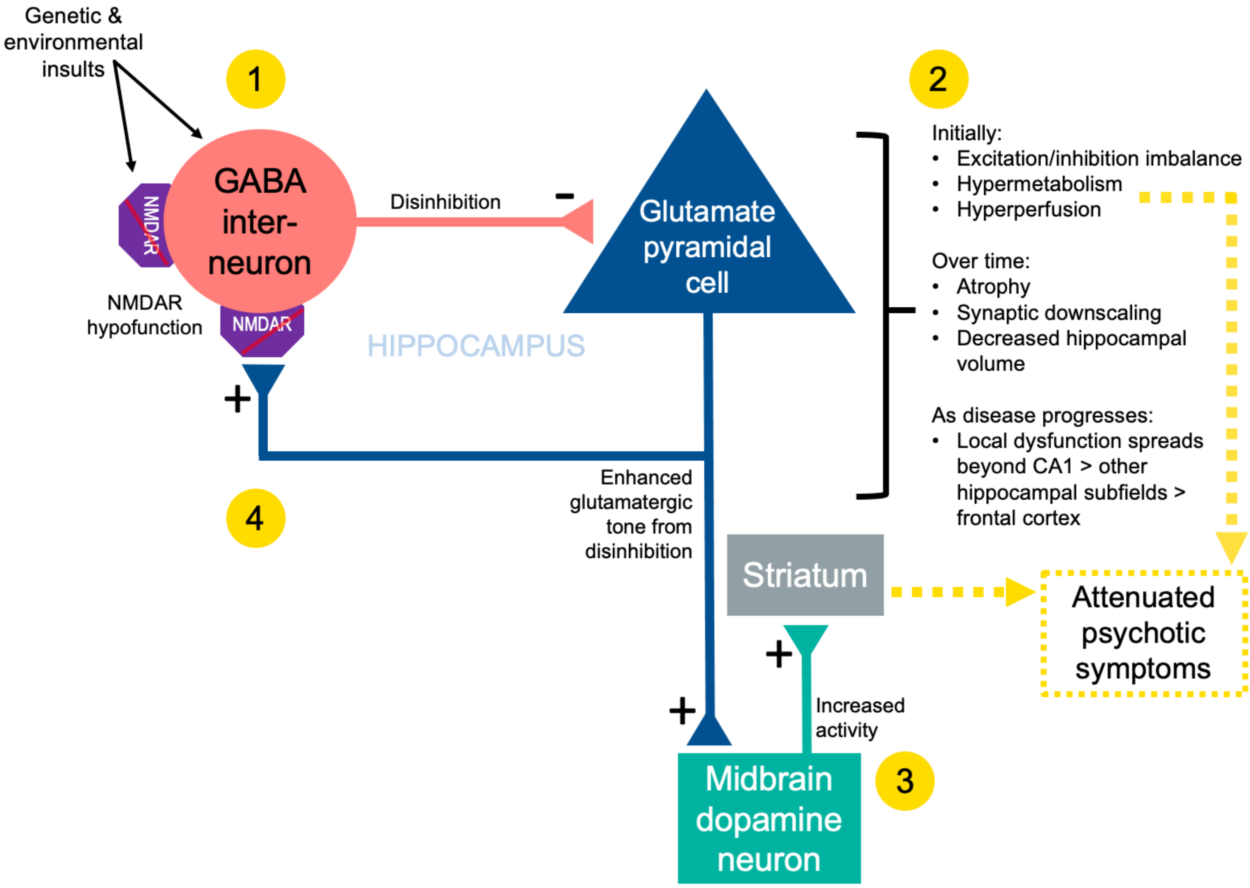

### SUPPLEMENTARY METHODS

#### MRI Acquisition and Image Processing

*Image Acquisition*

All scans were acquired (eyes-open) on a General Electric Signa HDx 3T MR system with an 8-channel coil at the Institute of Psychiatry, Psychology and Neuroscience, King's College London. For image registration both a high resolution T2-weighted Fast Spin Echo (FSE) image (TE= 54.58ms, TR= 4380ms, Flip angle= 90deg, FoV= 240, Matrix size= 320 x 320, slice thickness= 2mm, 72 spatial locations) and a high-resolution T1-weighted Spoiled Gradient Recalled (SPGR) image (TE= 2.85ms, TR= 6.98ms, Flip angle= 11deg, FoV= 260, Matrix size= 256 x 256, slice thickness= 1.2mm, 196 spatial locations) were acquired.

Resting Cerebral Blood Flow (CBF) was measured using 3D pseudo-Continuous Arterial Spin Labelling (CASL) scans acquired with a 3D Fast Spin Echo (FSE) spiral multi-shot readout, following a post-labelling delay of 1.5s. The spiral acquisition used a short (10ms) TE, and 8 spiral arms (interleaves) with 512 points in each arm. FSE TE= 32.26ms, TR = 5500ms. 64 slices of 3mm thickness were obtained and the in-plane FoV was 240×240mm. Three pairs of tagged-untagged images were collected. The whole ASL pulse sequence, including the acquisition of calibration images, was performed in 6:08min.

*Image Processing*

Data were preprocessed using FMRIB Software Library (FSL) 6.0.2 using the following procedure: (1) T1 and T2 images were skull-stripped and corresponding brain-only binary masks created; (2) original CBF images were coregistered to the T2 images and (3) multiplied by the binary T2 mask to create a skull-stripped CBF image in T2 space; (4) skull-stripped T2 was coregistered to skull-stripped T1; (5) skull-stripped T1 was first linearly coregistered to the MNI152 T1 2mm brain template, before non-linear registration (FNIRT) of the original T1 to MNI space; (6) original T2 images were registered to the MNI template (via T1 space) in a single concatenated step, using the T2-to-T1 transformation matrix (from step 4) and T1-to-MNI warp (from step 5); (7) skull-stripped CBF images (already in T2 space) were registered to the MNI template using the concatenated procedure in step 6; (8) normalised CBF images were spatially smoothed with a 6mm Gaussian kernel. The final voxel size was 2 x 2 x 2 mm. All images were visually inspected for preprocessing errors.

*Statistical Thresholds in SPM*

Statistical thresholds for exploratory wholebrain analyses (cluster-forming threshold: $p<.005$; cluster reported as significant at $p<.05$ using FWE cluster correction in SPM) were determined *a priori* based on previous work at our Institute investigating the effects of potential novel pharmacotherapies on rCBF in humans (Paloyelis *et al.*, 2016; Martins *et al.*, 2020b, 2022), including in our previous work in CHR patients (Davies *et al.*, 2019), and are standardly applied in ASL studies measuring rCBF (Joe *et al.*, 2006; Takeuchi *et al.*, 2011; Loggia *et al.*, 2013; Mutsaerts *et al.*, 2019; Martins *et al.*, 2020a; Nwokolo *et al.*, 2020).

### SUPPLEMENTARY RESULTS

#### CONSORT Details

The study was registered (ISRCTN46322781): https://doi.org/10.1186/ISRCTN46322781. Further details required for adherence to CONSORT (including recruitment periods, power calculations, randomisation and further blinding details, etc) can be found in the Supplementary Material of our previous publication in the same sample, where the study protocol is also appended (Bhattacharyya *et al.*, 2018a).

**FIGURE S2. CONSORT Flow Diagram (CHR patients)**

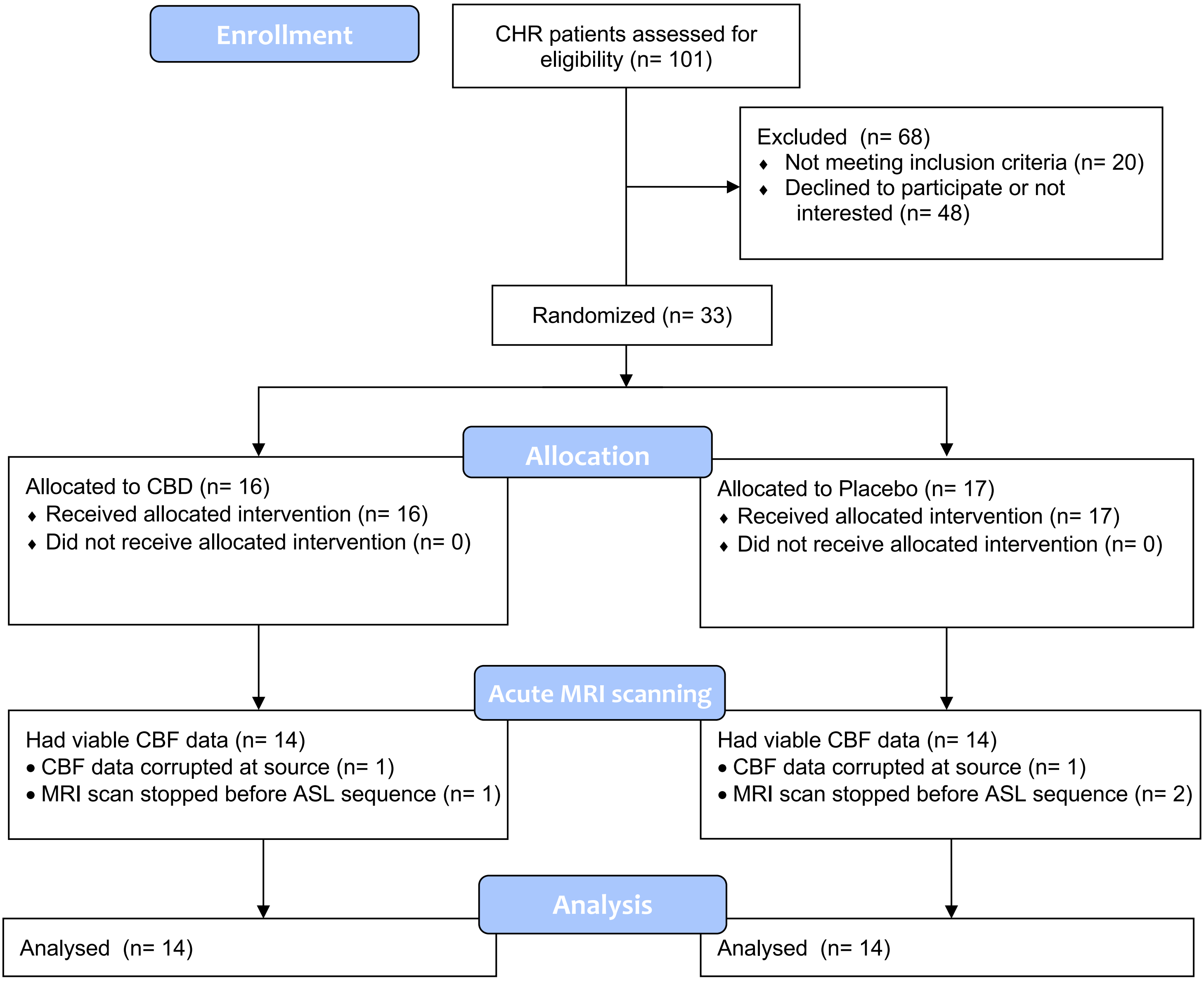

**FIGURE S3. Plot showing CBD plasma levels in CHR placebo and CBD groups**

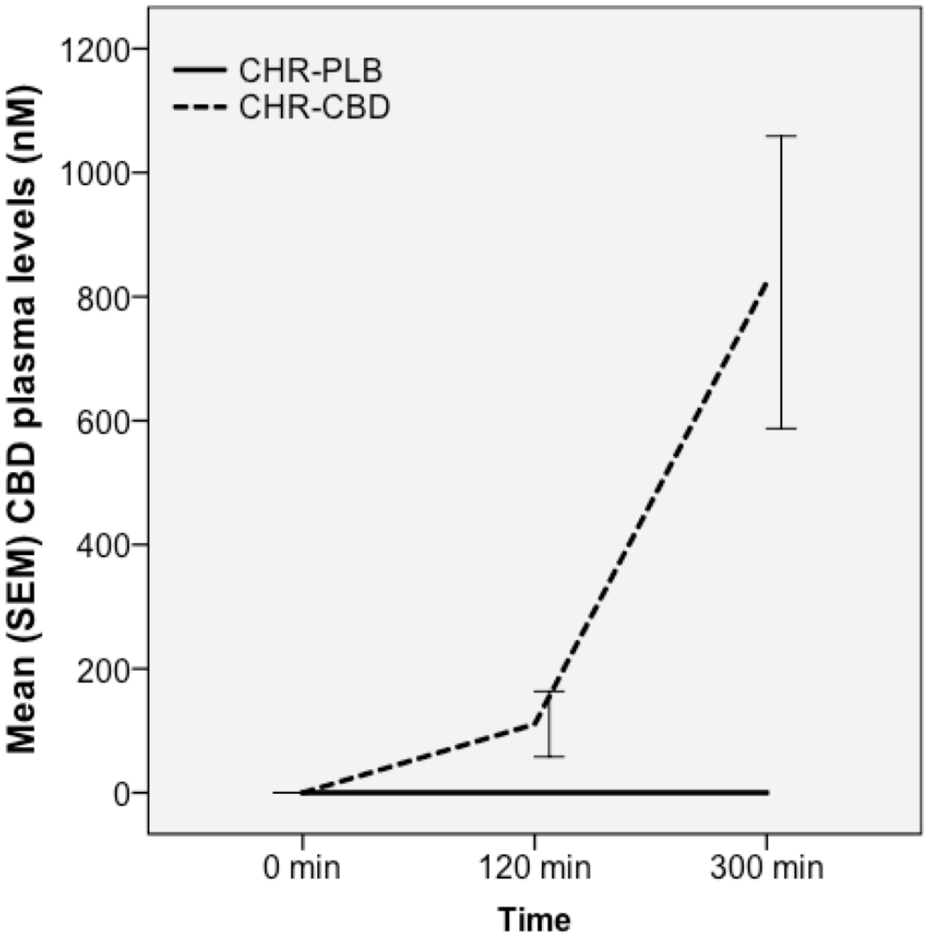

### Supplementary Wholebrain Analyses: SPM vs FSL Randomise

To test the robustness of the wholebrain findings, we re-ran our pairwise wholebrain analyses using two independent t-tests (controls vs placebo; placebo vs CBD) using *randomise* in FSL/6.0.1. De-meaned age, sex, years of education, smoking status and mean grey matter CBF per subject were included as covariates in the design matrix. The analysis used 5000 permutations and was restricted using a grey matter mask (thresholded at >.50). Cluster-based thresholding (threshold=2.3 due to modest sample size) was used, corrected for multiple comparisons by using the null distribution of the max (across the image) cluster size.

#### Healthy Control vs CHR Placebo

We found significantly higher CBF in a single (large, k=3660) cluster in the CHR placebo group vs healthy controls (cluster $p_{FWE}$=.025; see randomise output table below). In terms of anatomical location, the significant cluster found here using FSL's randomise (shown in red in the figure below) was almost identical to the significant clusters in our original analyses using SPM (shown in yellow in the figure below, superimposed on the FSL results [red]), with the addition of some further left cerebellar coverage with the FSL results. In terms of spatial extent, the FSL-derived clusters included slightly more voxels than the SPM results.

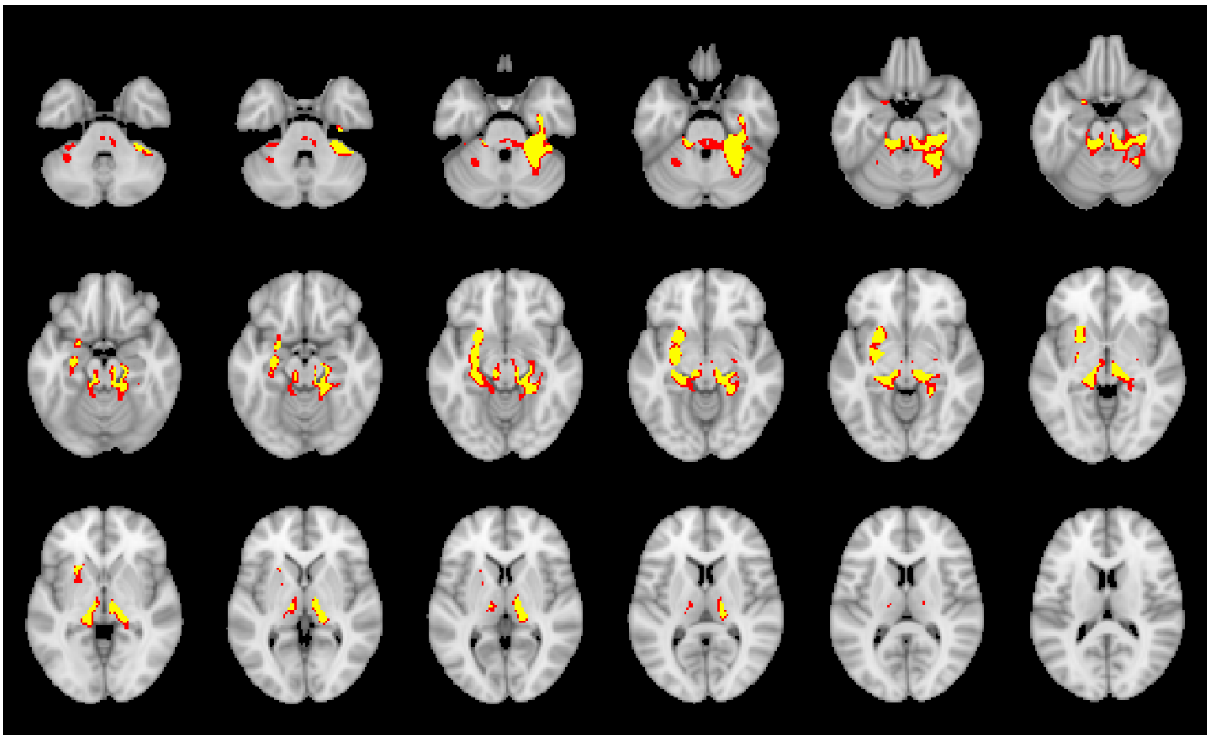

| Cluster Index | Voxels | 1-p-MAX | 1-p-MAX X (vox) | 1-p-MAX Y (vox) | 1-p-MAX Z (vox) | 1-p-COG X (vox) | 1-p-COG Y (vox) | 1-p-COG Z (vox) | COPE-MAX | COPE-MAX X (vox) | COPE-MAX Y (vox) | COPE-MAX Z (vox) | COPE-MEAN |
|---|---|---|---|---|---|---|---|---|---|---|---|---|---|
| **1** | 3660 | 0.975 | 61 | 41 | 13 | 43.1 | 49 | 27.5 | 4.98 | 58 | 68 | 32 | 2.9 |

**Placebo vs CBD**

For the CHR placebo vs CBD contrast, we did not observe any significant clusters using FSL's randomise. However, the cluster found using SPM (where CBD > placebo) was present at a relaxed statistical threshold (cluster $p_{FWE}$=.15; see randomise output table below). In terms of anatomical location, the (non-significant) cluster found using FSL's randomise (shown in red in the figure below) was almost identical to the significant cluster in our original analyses using SPM (shown in yellow in the figure below, superimposed on the FSL cluster [red]). In terms of spatial extent, the FSL-derived cluster included slightly more voxels than the SPM results, but note that this cluster was not significant in the FSL analysis. The reasons for the differential findings for this contrast in SPM vs FSL are unclear, but it is possible that the magnitude of the difference (SPM results: T(21)=4.51, $p_{FWE}$=.014) was not sufficiently large—combined with the modest sample size—to be significant with FSL's non-parametric statistical tests.

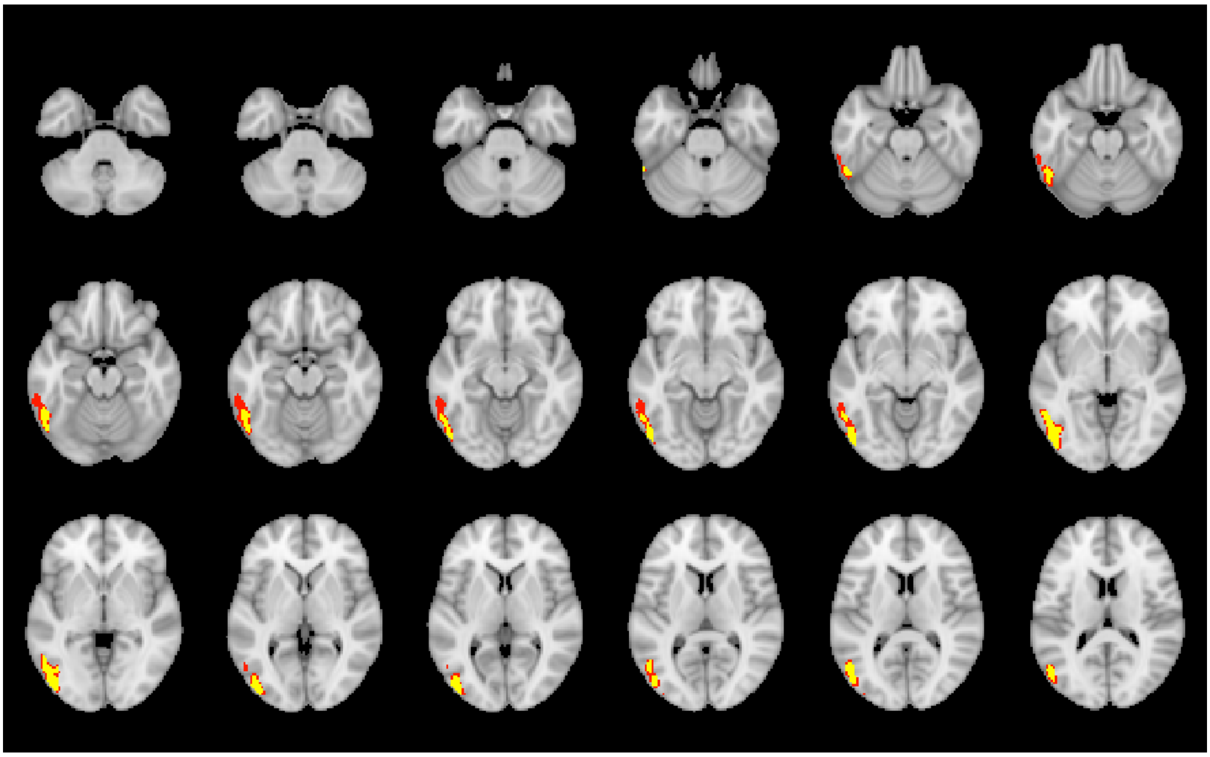

| Cluster Index | Voxels | 1-p-MAX | 1-p-MAX X (vox) | 1-p-MAX Y (vox) | 1-p-MAX Z (vox) | 1-p-COG X (vox) | 1-p-COG Y (vox) | 1-p-COG Z (vox) | COPE-MAX | COPE-MAX X (vox) | COPE-MAX Y (vox) | COPE-MAX Z (vox) | COPE-MEAN |
|---|---|---|---|---|---|---|---|---|---|---|---|---|---|
| **1** | 1206 | 0.854 | 72 | 38 | 22 | 70.6 | 30.9 | 33.2 | 4.4 | 67 | 23 | 39 | 2.83 |

**Exploratory Correlations between CAARMS and rCBF**

As we collected CAARMS data prior to drug administration and scanning, only baseline values from the CHR-placebo group could have been used in analyses with the imaging data, and given the modest sample size, we did not plan any *a priori* hypotheses along these lines. However, to explore whether hippocampal rCBF may be related to CAARMS scores, we correlated CAARMS positive symptoms (sum of the product of severity x frequency for each of the 4 positive symptoms) and total symptoms with rCBF values in the hippocampus (from the significant cluster from the linear trend/relationship analyses) in the placebo group. We found no significant correlation between rCBF and either CAARMS positive symptoms ($r=.065$, $p=.83$, $n=14$) or total symptoms ($r=-.073$, $p=.80$, $n=14$). This is consistent with two previous studies conducted at our institute, which found no significant association between attenuated positive symptom scores and elevated hippocampal rCBF in CHR patients (Allen *et al.*, 2016, 2018).

### SUPPLEMENTARY DISCUSSION

#### rCBF effects vs previous fMRI findings

Using the same patient and control sample, we previously demonstrated that CBD has effects on task-based BOLD haemodynamic readouts (Bhattacharyya *et al.*, 2018b; Wilson *et al.*, 2019; Davies *et al.*, 2020), finding the same commensurate pattern of placebo > CBD > controls (or vice versa) in mediotemporal regions during fear processing and verbal memory fMRI (Bhattacharyya *et al.*, 2018b; Davies *et al.*, 2020). CBF is intrinsically linked to BOLD responses via neurovascular coupling (Kim *et al.*, 2020), but its acquisition does not require the cognitive or other manipulation needed for task-based fMRI contrasts (Alsop *et al.*, 2015). As such, CBF can be used to index more proximal basal resting-state conditions (Alsop *et al.*, 2015). Moreover, it permits exploration of quantitative pharmacological effects across the brain without being spatially restricted to (and dependent on) the regions engaged by specific fMRI tasks. Capitalising on these advantages, our results extend previous knowledge by suggesting that CBD may also attenuate basal resting-state hippocampal activity in CHR patients. An interesting corollary of this finding is that in patients with early psychosis, hippocampal hyperperfusion has been directly associated with reduced hippocampal BOLD signal during fMRI scene processing (McHugo *et al.*, 2019). Increased basal perfusion combined with an attenuated stimulus-driven activation has also been observed in the amygdala in patients with schizophrenia (Pinkham *et al.*, 2015). These findings raise the possibility that elevated baseline activity (i.e. hyperperfusion) might be limiting effective recruitment during task performance (McHugo *et al.*, 2019). Although speculative, if CBD is indeed able to partially normalise basal hippocampal hyperperfusion this may, in turn, allow hippocampal circuitry to be recruited normally to meet mnemonic or other cognitive demands. Future CHR studies that combine perfusion imaging and memory fMRI (ideally with concomitant measures of performance), together with a CBD challenge, would allow investigation of this possibility.

#### Mechanisms

The molecular mechanisms by which CBD might have effects on hippocampal activity and rCBF remain unclear (Pertwee, 2008), but preclinical and *in vitro* work suggests that the general effects of CBD may be mediated by various mechanisms, including negative allosteric modulation of the CB1 receptor (Laprairie *et al.*, 2015). CB1 is highly expressed in hippocampus (Glass *et al.*, 1997) and CB1 agonism has been shown to disinhibit glutamatergic pyramidal neurons (Hájos *et al.*, 2000), an effect directly related to circuit-based models of psychosis (Fig S1) and which may be predicted to increase hippocampal blood flow (Lisman *et al.*, 2008; Knight *et al.*, 2022). By 'antagonising the agonists' of CB1 and impacting hippocampal endocannabinoid tone, it is possible that CBD modulates CBF through these direct receptor/circuit mechanisms. Further proposed mechanisms include inhibition of anandamide hydrolysis (Bisogno *et al.*, 2001) and actions on 5-HT1A (Russo *et al.*, 2005), vanilloid type 1 (Bisogno *et al.*, 2001) and GPR55 receptors (Ryberg *et al.*, 2007; Pertwee, 2008). Recent work has also implicated effects on the glutamate system (Gomes *et al.*, 2015; Linge *et al.*, 2016), which is of particular relevance to psychosis pathophysiology (Lodge and Grace, 2011; Howes *et al.*, 2015; Bossong *et al.*, 2019). On the human neuroimaging level, CBD modulates hippocampal glutamate in patients with early psychosis while concomitantly reducing psychotic symptoms (O'Neill *et al.*, 2021), and may partially ameliorate glutamatergic dysfunction in those at CHR (Davies *et al.*, 2023). CBD also alters glutamate and GABA in ASD and neurotypical individuals (Pretzsch *et al.*, 2019). The mechanisms underlying the effects of CBD on these neuroimaging parameters as well as on symptoms therefore remains an important avenue for future research.
